# Development of an implementation package for Asthma Medication Optimisation in the Emergency Department (AMEND) - an evidence, theory and person-based approach

**DOI:** 10.64898/2026.02.25.26346779

**Authors:** Imogen Skene, Ben Bloom, Jasmin Bassi, Anna De Simoni, Katy Pike, Chris Griffiths, Paul Pfeffer, Liz Steed

## Abstract

**Background:** Salbutamol is the most commonly prescribed inhaler for adults discharged from the Emergency Department (ED) with uncontrolled asthma. However newer options, e.g. Maintenance and Reliever Therapy (MART), are now recommended due to growing concerns over risks linked to salbutamol over-prescription. Transitioning to new inhalers requires support for both patients and healthcare professionals (HCP). This paper outlines how we developed an implementation package based on evidence, theory and the person-based approach to support asthma medication optimisation in the ED.

**Methods:** The purpose of this study was to use person-based intervention development methods in a three phase process: (1) understanding behaviour – collating and synthesising evidence from in-depth interviews with the target population (patients and HCPs); secondary deductive analysis using the theoretical domains framework (TDF) to understand barriers and facilitators to prescribing; developing guiding principles and logic model based on underlying theory (2) identifying behavioural content and implementation options - behaviour change techniques were selected and translated into intervention content (3) intervention materials developed and refined with input of stakeholders.

**Results:** We identified modifiable target behaviours for HCPs to support guideline-based care in the ED. These included identifying eligible patients, communicating rationale, providing patients with inhaler and resources, and communicating changes to primary care.

Key theoretical domains included knowledge, skills, addressing beliefs about consequences, and targeting professional role perceptions. These domains were targeted through a clinical decision aid and training materials for ED HCP, template discharge summary for primary care, and visual and written materials for patients. Minor refinements were made based on stakeholder feedback (six ED doctor think-aloud interviews; two patient workshops with eight and five participants respectively; and 12 survey responses to final draft of video).

**Conclusion:** We developed an intervention grounded in theory, evidence, and stakeholder feedback aimed at promoting and supporting delivery of guideline recommendation.

**KEY MESSAGES:** *What is already known:* ED visits represent teachable moments where patients may be receptive to optimising asthma medication, yet guideline-concordant prescribing is often limited in fast-paced settings.

*What this study adds:* This study describes AMEND, a behaviourally informed implementation package including a clinical decision aid, HCP training, and patient-facing materials, designed to support initiation of Maintenance and Reliever Therapy (MART) at discharge. Using stakeholder-informed, iterative development, the package was feasible to integrate into routine ED workflows.

*How this study might affect research, practice or policy:* Findings highlight how theory-driven interventions can translate guidelines into practice, improve adherence, and potentially reduce repeat ED attendances.

## Introduction

Asthma management has traditionally relied upon the use of separate preventer (inhaled corticosteroids – ICS) and reliever (short-acting beta-agonist - SABA) inhalers. However, many people underuse their preventer and overuse their reliever, a pattern associated with increased risk of hospital admission and poor asthma outcomes(1, 2). Emergency Departments (EDs) often see patients during periods of poor asthma control and therefore represent an important opportunity to identify suboptimal management and initiate safer treatment strategies.

Recent national and international guidelines now recommend maintenance and reliever therapy inhaler (MART) – using a low-dose ICS/formoterol inhaler taken both regularly and as needed - instead of SABA (3). This approach ensures patients receive ICS whenever they use a reliever, even when adherence to scheduled preventer therapy is poor, leading to improved asthma control (4). These guidelines represent a significant change in UK clinical practice.

Implementation of MART has focused on primary care (3, 5), yet those with uncontrolled asthma often present directly to the ED. Follow up after hospital attendance is often limited, with a minority of adults receiving a timely medication review, with patients from black and ethnic minority groups least likely to receive any post hospitalisation care (6). Moreover, ED attendance has been associated with increased adherence to recommendations (7) suggesting the ED as a potentially powerful, but underutilised setting for optimising asthma treatment and addressing health inequalities.

Intervention delivery in the ED is challenging due to time pressures, completing priorities, and the acute nature of ED practice. Although ED based interventions show potential to improve longer-term asthma outcomes (8), successful translation of interventions into practice has been limited (9). Updated Medical Research Council (MRC) guidance emphasises the importance of considering implementation processes and context during intervention development to improve uptake and sustainability (9). While asthma guidelines (3) and discharge bundles (10) have been updated, the practical guidance on implementation in ED settings remains limited.

Implementation initiatives are more effective when developed with key stakeholders including patients and healthcare professionals (HCP) (11, 12, 13, 14). The person-based approach (PBA) prioritises understanding users’ perspectives, to ensure interventions are acceptable and feasible (11). Qualitative studies indicate that patients attending the ED with acute asthma are receptive to medication optimisation when clearly explained and supported by information to take home (15), and ED HCP are willing to prescribe improved therapy at discharge when supported by simple processes and training (16).

This evidence indicates readiness to support an ED based implementation package to optimise asthma medication at discharge (15, 16). We hypothesise that optimising asthma mediation at the point of ED discharge could improve asthma outcomes and reduce readmissions. To address this, we developed an implementation package to support asthma medication optimisation in the ED

## METHODS

We have followed the guidance for reporting intervention development studies in health research (GUIDED) in this paper (17).

### Study Design

This study used a systematic approach to develop an implementation package aimed at asthma medication optimisation in the ED (AMEND). The development process was informed by the Medical Research Council (MRC) framework for complex interventions, but drew specifically on the Behaviour Change Wheel (BCW) and the Person-based approach (PBA) to guide the content and delivery strategy (9, 11, 18). PBA describes iterative methods of planning, optimising, evaluating and implementing behavioural health interventions. It is typically based on mixed methods research where users’ views and experiences are central to ensure the intervention is engaging and persuasive (19). The implementation package was grounded in theory through a secondary analysis of qualitative data using the Theoretical Domains Framework (TDF) (20, 21) and application of the Perceptions and Practicalities Approach (PAPA) (22). The TDF was used to identify theoretical constructs influencing behaviour and has been applied effectively in similar ED based implementation studies (23, 24). PAPA was applied to distinguish between perceptual barriers (e.g. beliefs about asthma control or treatment necessity) and practical barriers (e.g. inhaler availability, ED constraints). This analysis informed the content and delivery strategy of AMEND ensuring that the intervention address both motivational and practical determinants of behaviours, supporting feasible and sustainable implementation.

An AMEND development group was formed to provide iterative feedback from people with asthma, public contributors, HCPs, and a multi-disciplinary expert working group, with expertise in implementation science, behaviour change, complex intervention development, respiratory, emergency and primary care, and qualitative methodologies. Figure 1 summarises the phases and key steps in the development process.

**Figure 1:**
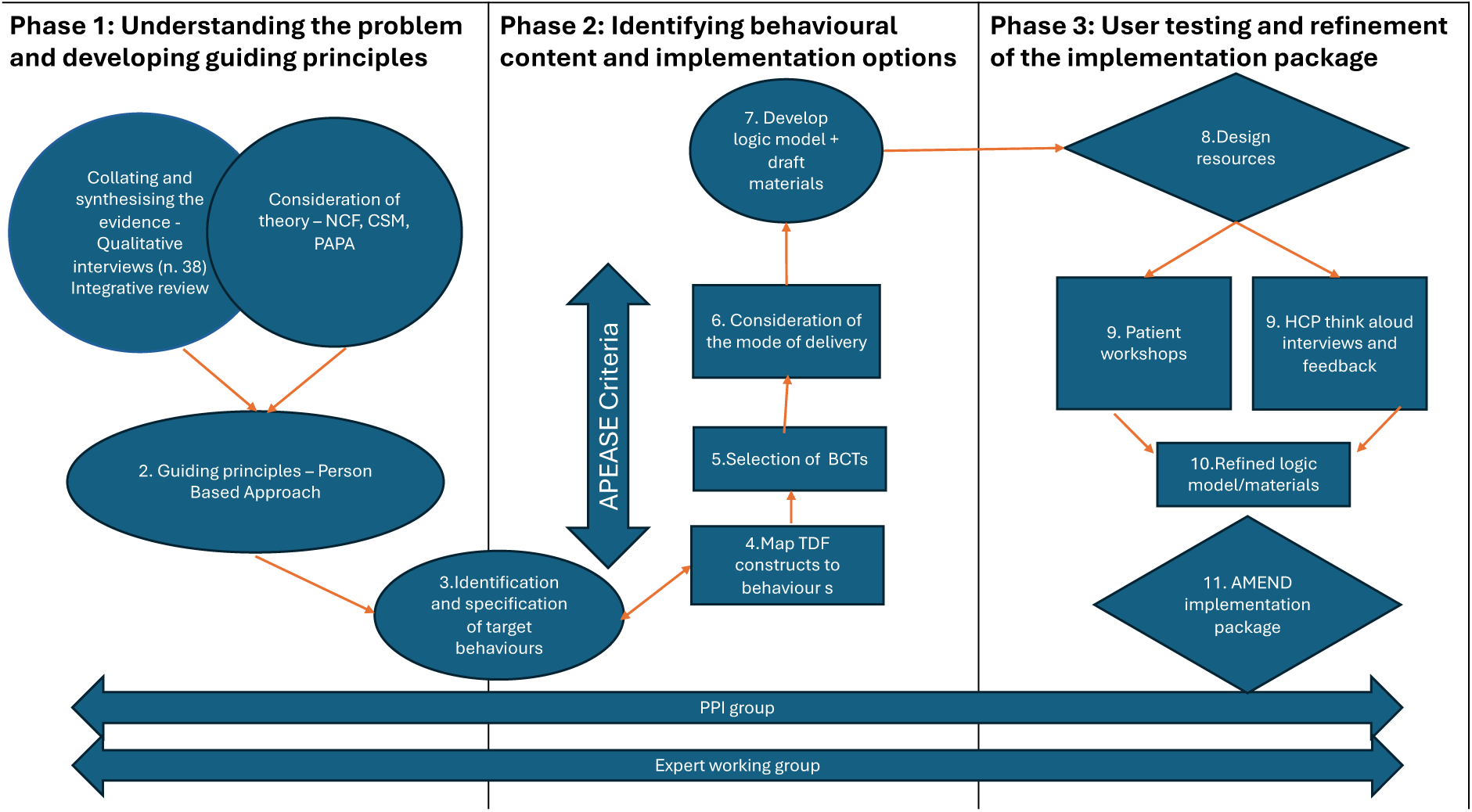
The three phases of the development of the AMEND implementation package

### Phase 1: Understanding the Problem and Developing Guiding Principles

In line with MRC guidance we reviewed the literature (8) and conducted qualitative studies (15, 16) to explore initial need and understand the problem. We then developed ‘guiding principles’ to specify ‘what’ the design objectives must be, and ‘how’ they may be achieved (25). We used a triangulation method to identify key recurring themes across the data sources or those given high priority by respondents. Initial guiding principles were discussed with the intervention development group before finalisation.

### Phase 2: Identifying behaviour, content and implementation options

This phase involved several key steps each performed for patient, HCP and organisational behaviours

1) Identifying target behaviours. In line with the BCW the behavioural pathway was specified. Behaviours were then rated according to APEASE criteria (Acceptability, Practicability, Effectiveness, Affordability, Side-effects, and Equity) and final target behaviours agreed through discussion with the expert working group and patient and public involvement (PPI) contributors.

2) Mapping TDF constructs to target behaviours

a) Using methods described by Atkins (2017) (26) and the qualitative data previously collected from patients and HCPs (15, 16) deductive coding was carried out
b) Domains were considered relevant if there was data that reflected the domain and it related to a target behaviour
c) Within domains, key belief statements were summarised to interrogate the most important constructs for targeting.

3) Selecting behaviour change techniques (BCTs)

Once the key TDF domains and constructs were identified, we mapped BCTs that could enable change. For example, to address skills gaps, BCTs including instructions on how to perform a task and behavioural practice were selected. At this point we brought in additional theoretical knowledge where appropriate e.g. when targeting the domain beliefs about consequences we drew on PAPA to consider concerns around necessity versus worries related to medication.

4) Determining mode of delivery

We then determined the most appropriate mode of delivery for each BCT. This involved selecting delivery formats which would be practical for delivery within a fast-paced environment.

5) Developing the logic model

A logic model was developed, in line with MRC guidance, to visually map the hypothesised theoretical mechanism of the intervention (27, 28).

6) Developing materials

We then proceeded to develop the prototype materials for the implementation package.

### Phase 3 – User testing and Refinement

User testing and refinement of the materials was iterative but is presented as a final step here for ease, see Figure 2. This involved suggesting and agreeing content, assessing useability and identifying potential barriers and facilitators. User testing with stakeholders was conducted separately for people with lived experience of asthma and HCPs. Participants received a shopping voucher as a thank you for their time. Feedback was prioritised using the MoSCoW “Must have”, “should have”, “could have” and “won’t have” method to guide revisions based on their importance(29).

**Figure 2:**
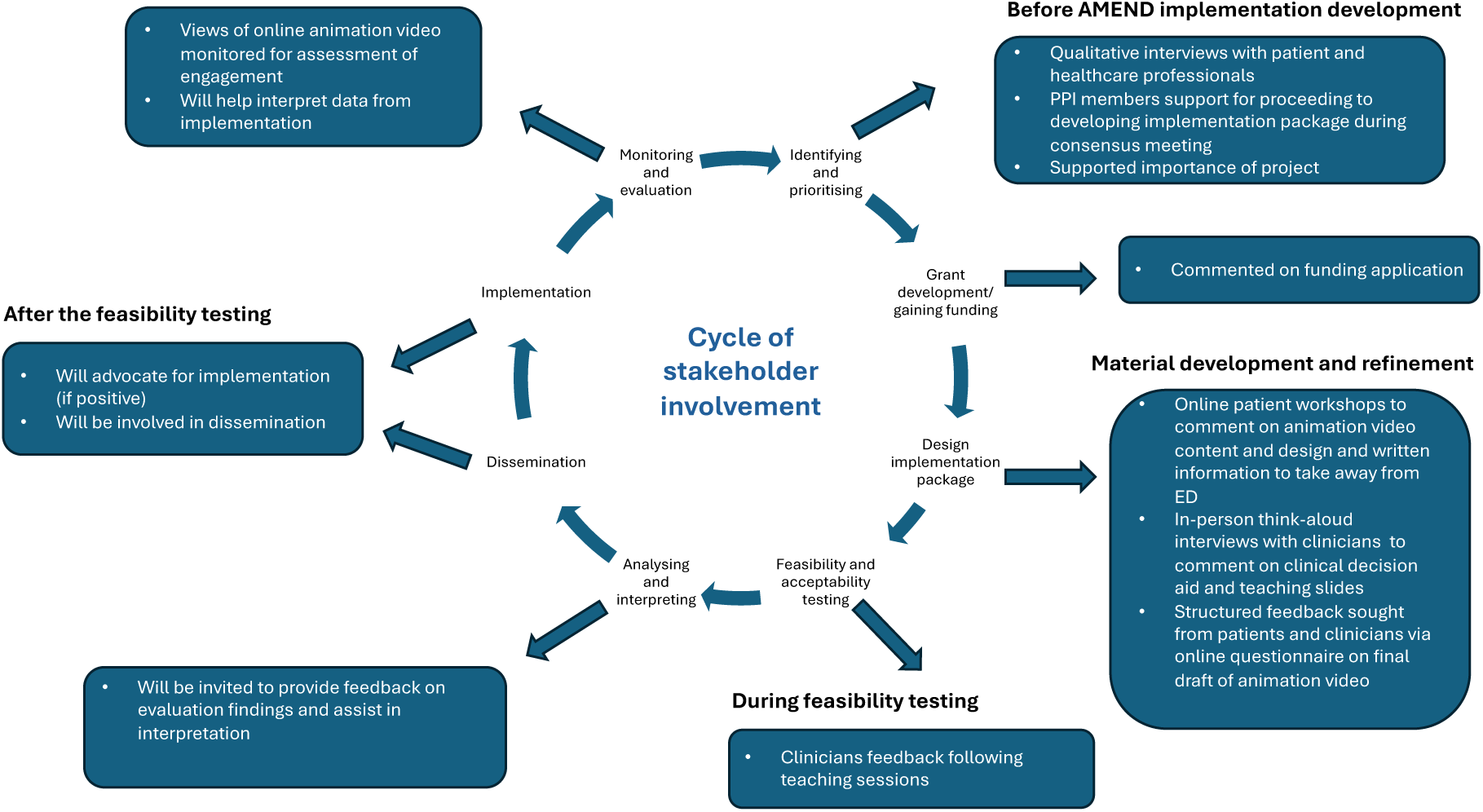
Stakeholder involvement in the AMEND project>

Patient facing materials: Lived experience perspectives were gathered through online workshops. Participants were recruited via outreach to the research teams clinical and professional networks, respiratory patient groups (Asthma + Lung UK, Respiratory Insights) and our PPI group. Eligible participants were adults with a history of attending the ED for asthma related care. Participants were provided with information about the quality improvement project in advance and consent was by participation. Workshops included brief presentations of draft materials followed by facilitated discussion. Field notes and informal participant feedback were collected during the sessions and used to inform refinement of the materials.

HCP facing materials: HCP testing involved “think aloud” interviews with ED HCPs. This method enables participants to verbalise their thoughts while interacting with the materials, offering insights into usability and cognitive processes (11). ED doctors from the trust where the implementation package would be piloted were invited to participate via email. We sought a sample of both resident doctors and consultants to capture a range of perspectives.

Each session began with a scripted introduction. Participants were provided with information about the quality improvement project and gave verbal consent. Participants were informed that participation was voluntary, they could withdraw at any time and that the interview would be audio-recorded for analysis purposes and used to inform the refinement of the materials. Participants then engaged with the materials using two example patient cases while verbalising their thoughts. Open-ended questions at the end of each session elicited feedback on usability, acceptability, and design. Following initial refinement, additional ongoing structured feedback was collected using online survey methods (Microsoft Teams forms), to further refine materials. Participants were provided with information about the purpose of the feedback and how their responses would be used. Participation was voluntary, and consent was by submission of the online form.

### Ethics and Consent to Participate

The qualitative research in phase 1 and 2 was conducted in accordance with the Declaration of Helsinki and ethical approval was obtained from Camden and Kings Cross Research Ethics Committee HRA Approval [REC Reference 21/LO/0665]. The study received NHS governance approval from Barts Health NHS Trust. All participants provided written informed consent.

Phase 3, including the user testing and refinement were conducted as part of a quality improvement project, registered at Barts Health NHS Trust, to design the implementation package, deliver and evaluate its acceptability. This did not require separate ethical approval.

All participants in the think aloud interviews provided verbal informed consent. Participants of online workshops and structured survey feedback were informed that their participation was voluntary, that they could withdraw at any time and that all data would be treated confidentially; participation was considered as consent

### Patient and public involvement

The AUKCAR and Barts Health NHS Trust ED PPI group advised on the design of the primary qualitative interviews, ensuring the approach and timing was suitable to reach patients in the ED. At each stage of AMEND development the results were presented to AUKCAR PPI members to facilitate interpretation and to refine and help shape the final intervention materials.

## RESULTS

### Overview

The AMEND implementation package was developed through a structured, multi-phase process to address barriers to optimal asthma management at ED discharge.

### Phase 1: Understanding the behaviour

Table 1 presents the guiding principles and confirmed acceptability of an intervention in ED providing specific design considerations were made.

**Table 1:**
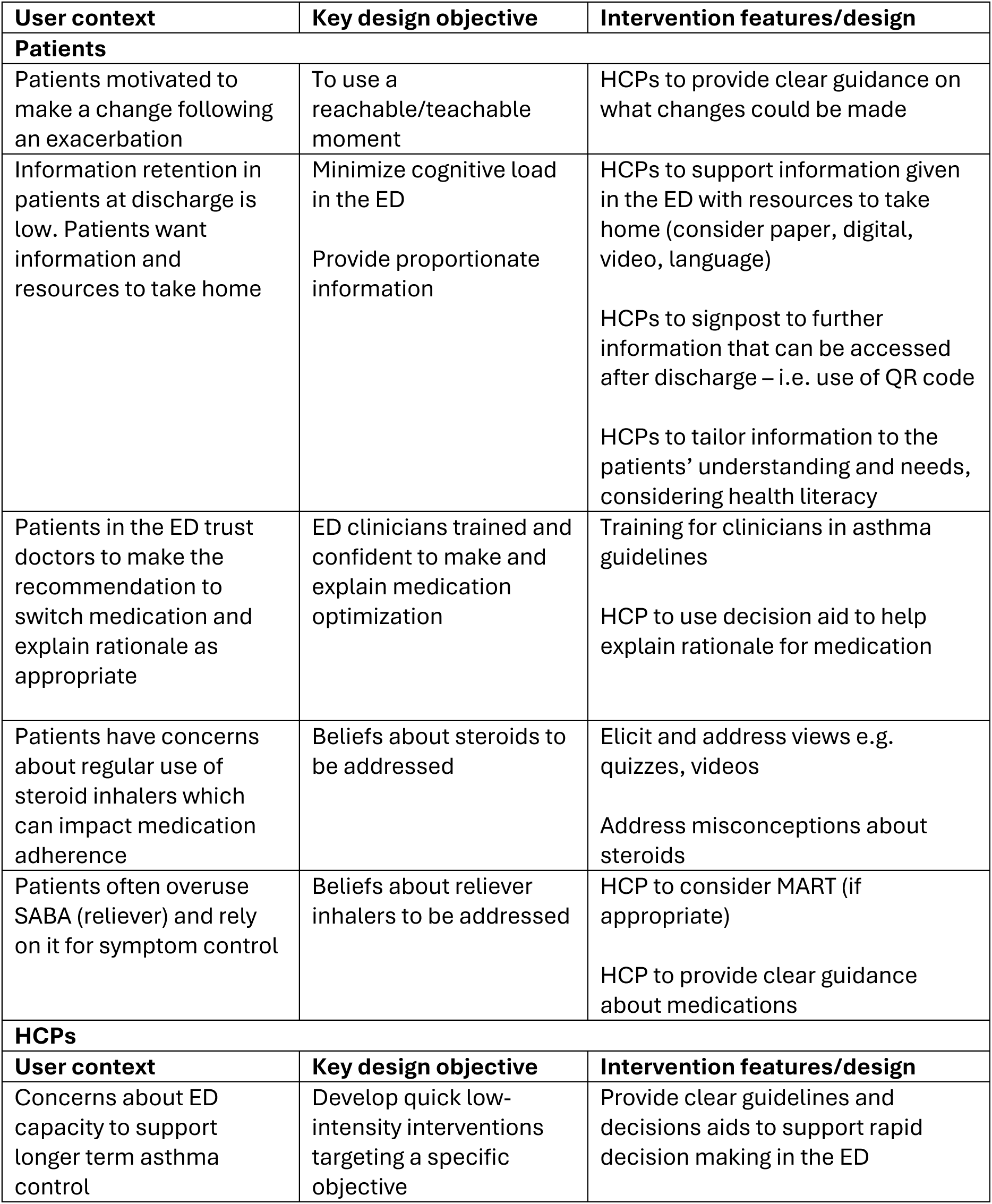
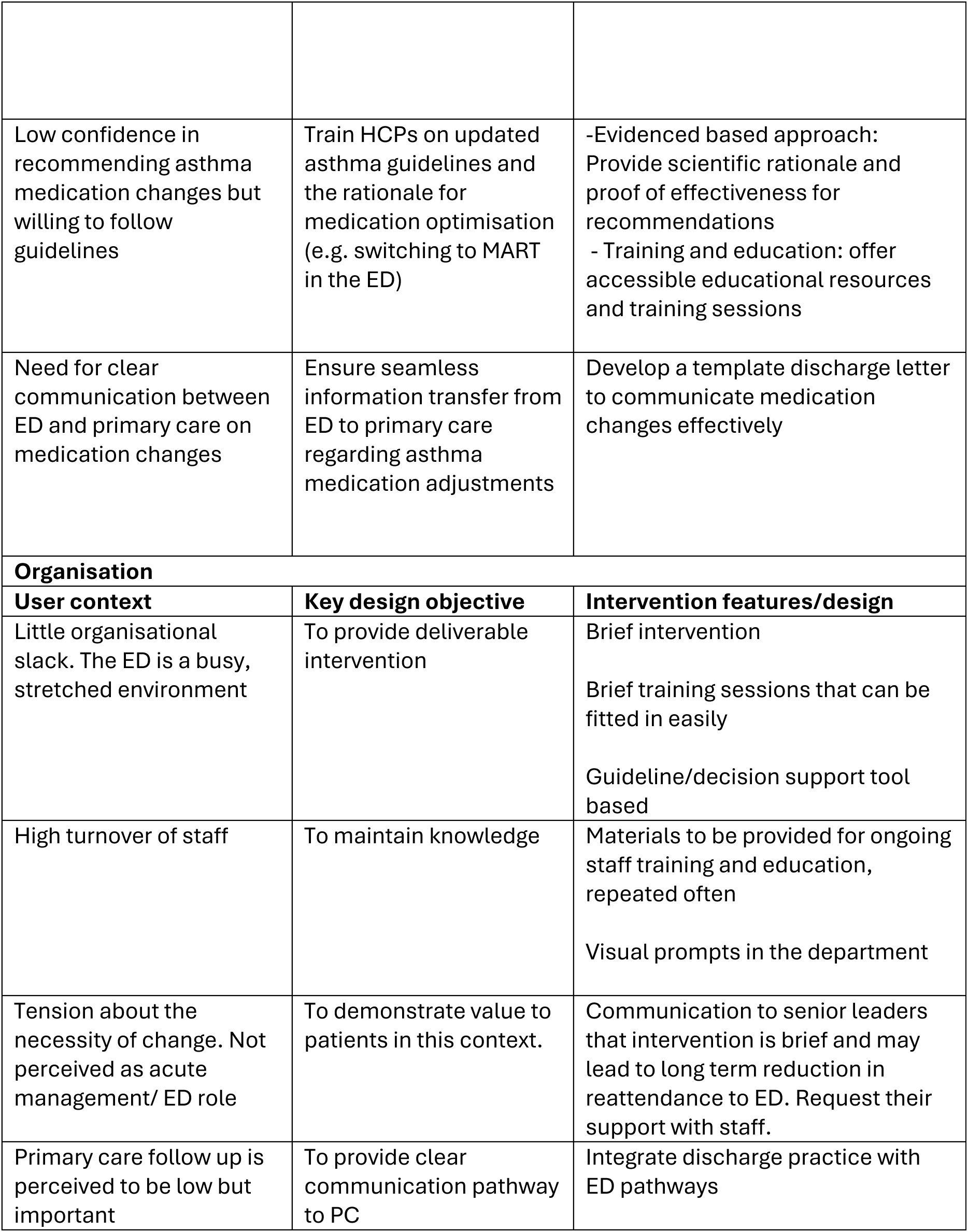
The guiding principles for the development of the AMEND implementation package.

| User context | Key design objective | Intervention features/design |
| --- | --- | --- |
| **Patients** |  |  |
| Patients motivated to make a change following an exacerbation | To use a reachable/teachable moment | HCPs to provide clear guidance on what changes could be made |
| Information retention in patients at discharge is low. Patients want information and resources to take home | Minimize cognitive load in the ED<br>Provide proportionate information | HCPs to support information given in the ED with resources to take home (consider paper, digital, video, language)<br>HCPs to signpost to further information that can be accessed after discharge – i.e. use of QR code<br>HCPs to tailor information to the patients' understanding and needs, considering health literacy |
| Patients in the ED trust doctors to make the recommendation to switch medication and explain rationale as appropriate | ED clinicians trained and confident to make and explain medication optimization | Training for clinicians in asthma guidelines<br>HCP to use decision aid to help explain rationale for medication |
| Patients have concerns about regular use of steroid inhalers which can impact medication adherence | Beliefs about steroids to be addressed | Elicit and address views e.g. quizzes, videos<br>Address misconceptions about steroids |
| Patients often overuse SABA (reliever) and rely on it for symptom control | Beliefs about reliever inhalers to be addressed | HCP to consider MART (if appropriate)<br>HCP to provide clear guidance about medications |
| **HCPs** |  |  |
| User context | Key design objective | Intervention features/design |
| Concerns about ED capacity to support longer term asthma control | Develop quick low-intensity interventions targeting a specific objective | Provide clear guidelines and decisions aids to support rapid decision making in the ED |
| Low confidence in recommending asthma medication changes but willing to follow guidelines | Train HCPs on updated asthma guidelines and the rationale for medication optimisation (e.g. switching to MART in the ED) | -Evidenced based approach: Provide scientific rationale and proof of effectiveness for recommendations<br>- Training and education: offer accessible educational resources and training sessions |
| Need for clear communication between ED and primary care on medication changes | Ensure seamless information transfer from ED to primary care regarding asthma medication adjustments | Develop a template discharge letter to communicate medication changes effectively |
| **Organisation** |  |  |
| **User context** | **Key design objective** | **Intervention features/design** |
| Little organisational slack. The ED is a busy, stretched environment | To provide deliverable intervention | Brief intervention<br>Brief training sessions that can be fitted in easily<br>Guideline/decision support tool based |
| High turnover of staff | To maintain knowledge | Materials to be provided for ongoing staff training and education, repeated often<br>Visual prompts in the department |
| Tension about the necessity of change. Not perceived as acute management/ ED role | To demonstrate value to patients in this context. | Communication to senior leaders that intervention is brief and may lead to long term reduction in reattendance to ED. Request their support with staff. |
| Primary care follow up is perceived to be low but important | To provide clear communication pathway to PC | Integrate discharge practice with ED pathways |

### Phase 2: Identifying content and implementation options

Five target behaviours were identified for the implementation package for optimising inhalers and medication change by the clinician which were:

1. Identify suitable patient
2. Identify correct inhaler
3. Communicate rationale and agree change with patient
4. Provide patient with inhaler and resources
5. Communicate change and follow up request to primary care

To effectively optimise asthma inhalers, it was recognised that an implementation package primarily targeting HCP level behaviours was required. HCPs behaviours in the ED most commonly represent the first and precipitating step in the causal chain and hence have spillover effect. Patient behaviour, such as their engagement with both the HCP and the materials developed is recognised as essential for successful implementation. However, as the patient behaviours would typically take place following clinician initiation (i.e. discussing MART inhaler for symptom relief), for simplification the implementation package is presented with a primary focus on HCP behaviour.

Table 2 reports full behaviour specification as well as TDF and BCT mapping for the target behaviours. Several key determinants were identified for the behaviours and related to established behaviour change theories, which were used for greater depth of theoretical understanding. For example:

- The Common Sense Model of illness self-regulation (CSM) (30) highlights beliefs about the severity and time-line of asthma, which influence adherence
- The Necessity-Concerns Framework (NCF) (31) clarifies concerns about inhaler necessity versus fears of side effects from long term steroid inhaler use
- The Perceptions and Practicalities Approach (PAPA) (22) addresses practical barriers such as difficulty using inhaler correctly.

**Table 2.** Target behaviours identified for change.

| Target behavior - What needs to be done by ED clinician | Where | When | Mapped TDF domains from barriers identified | BCT Applied | Implementation strategy |
| --- | --- | --- | --- | --- | --- |
| Identify suitable patient | All urgent and emergency care environments – using patient history, observations and electronic records | During initial assessment or discharge planning | **Knowledge** - Limited awareness of MART and new guideline criteria for switching inhalers<br>**Social/professional role and identity** - Switching patients long-term asthma medication is not always perceived as an ED responsibility<br>**Memory, attention and decision process** – Remembering criteria in a busy environment can be difficult | 4.1 Instruction on how to perform behaviours<br>6.3 Information about others approval<br>7.1 Prompts/ cues | Decision aid<br>Training package<br>Visual prompts |
| Identify correct inhaler | At staff workstations with access to electronic guidelines | After patient assessed and deemed suitable for inhaler change | **Knowledge** – Uncertainty about which inhaler to prescribe<br>**Beliefs about capability** - Lacked confidence in knowledge and skills to optimise inhalers or recommend a switch to MART | 5.1 Information about health consequences<br>4.1 Instruction / | Printed guideline flowchart<br>Decision aid<br>Training package |
|  |  |  | **Beliefs about consequences** – Concerns about prescribing outside usual ED scope | Demonstrat<br>ation of<br>how to<br>perform a<br>behavior<br>6.3<br>Informatio<br>n about<br>others<br>approval |  |
| Communi<br>cate<br>rationale<br>and<br>agree<br>change<br>with<br>patient | At the<br>bedside /<br>cubicle,<br>where a<br>private<br>conversatio<br>n can occur | During patient<br>consultation,<br>once inhaler<br>choice has<br>been made,<br>before<br>prescribing or<br>dispensing | **Skills** – Limited confidence in explaining rationale<br>**Social/professional role and identity** – Seen as time consuming in the ED setting and considered a primary care responsibility | 6.1<br>Modelling/<br>demonstra<br>tion<br>8.1<br>Behavioral<br>rehearsal<br>5.1 Informa<br>tion about<br>health<br>consequ<br>ences | Decision<br>aid<br>Training<br>package<br>Patient<br>video<br>Take-<br>home<br>leaflet |
| Provide<br>patients<br>with<br>inhaler<br>and<br>resource<br>s | At the<br>bedside /<br>cubicle,<br>with access<br>to inhaler,<br>spacer (if<br>needed),<br>and printed<br>information<br>sheet with<br>QR codes<br>for video | At point of<br>discharge,<br>after patient<br>is clinically<br>stable | **Environmental context and resources** - new inhaler and printed leaflets need to be available in the ED<br>**Memory, attention and decision process** – Clinicians may forget to provide resources at discharge<br>**Beliefs about consequences** – Perception that inhaler change is optional and will be reviewed later in primary care after discharge<br>**Skills** - Low confidence in giving advice on inhalers technique | 12.5 Add<br>object to<br>the environ<br>ment<br>7.1<br>Prompts/c<br>ues | Video<br>Take-<br>home<br>leaflet |
| Communicate change and follow up request to primary care | At staff workstation using electronic discharge summary | Immediately after discharge decision and patient education | **Environmental context and resources** - GPs required notification of inhaler changes for follow up<br>**Beliefs about consequences** – Clinicians recognized the importance of informed GP for safe follow up<br>**Social/professional role and identity** – Responsibility for GP follow up shared with patients' responsibility for making follow up appointment, but ED clinicians to provide clear discharge instructions. | 1.4 Action planning.<br>7.1 Prompting | Template discharge text |

Implementation package resources were drafted for refining in the next phase. The AMEND proposed logic model is presented in Figure 3.

**Figure 3:**
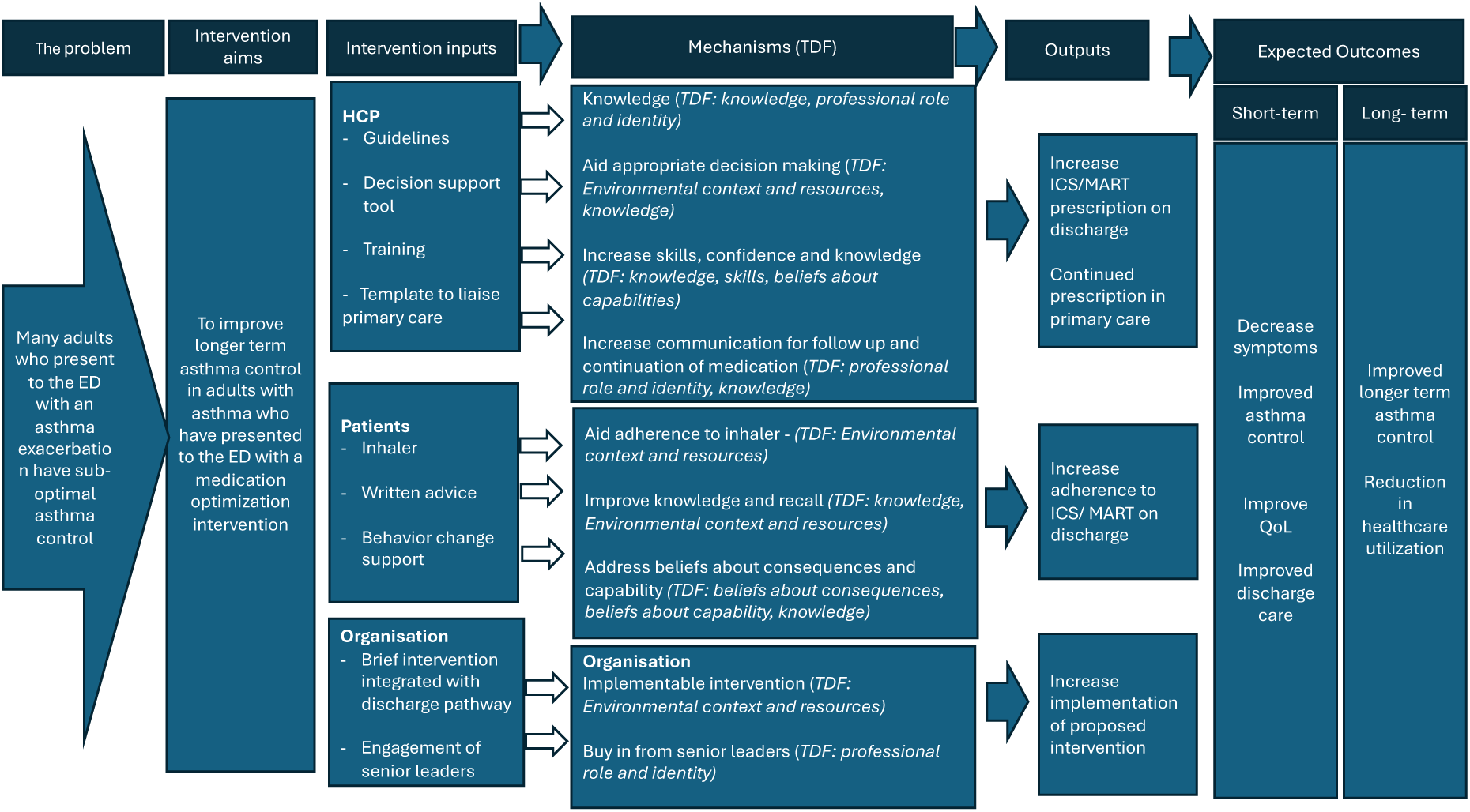
Logic model

### Phase 3 – User testing

#### Patient facing materials – animation video and written resources to take away

Two online workshops to gain lived experience perspectives were held in March 2024 and September 2024. Thirteen participants with a history of ED presentations for asthma (8 in workshop 1, 5 in workshop 2) participated. The majority had attended the ED in the last year (8/13), and 8/13 reported prior experience of using a MART inhaler. Feedback from the first workshop based on an animation storyboard highlighted

- the need to simplify messaging in the patient facing animation,
- avoidance of medical jargon
- stronger communication of the importance of ICS and the risks of SABA overuse
- additional resources to reinforce messages post-discharge.

In response, the intervention was revised - see Table 3 for changes.

**Table 3.** Example issues identified from the stakeholder engagement and changes implemented to address these.

| **Intervention aspect** | **Key feedback** | **Suggested change** | **Reason for change** | **Agreed change** | **Priority (MoScoW\*)** |
| --- | --- | --- | --- | --- | --- |
| Clinical decision aid – page 1 flow diagram | Clinicians may continue prescribing salbutamol alongside MART; flow requires clearer guidance. | Add warning sign and box: “Do not also prescribe a salbutamol inhaler” | Prevents unsafe duplicate prescribing; aligns with expert feedback. | Warning box added to flow diagram. | M |
| Clinical decision aid – page 1 flow diagram | Include a step in the flow diagram to check patient adherence before switching therapy. | Add adherence check step to flow diagram | Encourages guideline-concordant prescribing. | Not implemented; prioritized simplicity of flow chart. | C |
| Clinical decision aid – inhaler visuals | Decision aid does not show spacer with MDI inhalers | Add image of spacer to inhaler recommendations | Improves usability and reinforced correct behaviours- Resonates with views of expert group. | Spacer images added | S |
| Patients take home resources | Patients felt written information to take away was important | Provide written information and QR codes | Supports retention and adherence after discharge | Leaflet with QR codes provided | M |
| Patient animation video | Participants disliked idea of a quiz or questions incorporated as it would feel like a test and felt had the potential to be judged by doctors | Script adapted for information key messaging only | Supports engagement without patient anxiety | Quiz removed, key points emphasised | M |
| Patient animation video | Participants felt that well controlled asthma doesn't have symptoms needed to be emphasized | Repeat during video | Reinforced education and self-management understanding | Moved to opening and closing of video | M |
| Template discharge text | Clear patient facing discharge summary in decision aid is helpful to standardize information – would be helpful if auto populated on electronic patient record | Develop pre-populated discharge template on electronic patient record | Improves consistency and efficiency | Template discharge summary finalized and added to electronic patient record | M |
| HCP training | Concerns that the training is too long and would benefit from short version which could be played at handovers | Provide short, pre-recorded video | Improves accessibility and supports flexible learning. Improves reach where in-person teaching is less feasible. | Video and in person sessions implemented | S |
| HCP training | Concerns about time pressure and stock availability in the ED | Streamline processes and ensure MART inhalers in TTA cupboard | Addresses real world feasibility barriers | Added to implementation plan | M |
| **MoScoW= M** = “Must have”; S= “should have”; C= “could have”; and W = “won’t have” |  |  |  |  |  |

The second workshop reviewed the revised materials, using the storyboard visuals for the animation video. Participants confirmed acceptability with minor amendments to wording and visuals. No further substantial changes were suggested.

The full animation video was subsequently shared via email with a wider group for structured feedback. Twelve participants responded, including patients (n=4), PPI contributors (n=2), HCPs (emergency medicine doctor n=1, respiratory doctor n=4, nurses or allied health professionals n=2). Seven reported a history of asthma, and all had English as their first language. Survey feedback confirmed that the animation was clear, relevant, and acceptable to both patients and healthcare professionals, with no new major concerns raised.

*“Really like it - concise, clear and gets the message across” - Patient feedback via feedback form*

#### ED HCP facing materials: Clinical decision aid and teaching slides

HCP Think-Aloud Interviews: Six ED doctors participated in think-aloud interviews focused on the decision aids (3 male, 3 female), including consultants (n=2), ST4–6 trainees (n=1), ST1–3 trainees (n=2), and one FY2 doctor. All were emergency medicine doctors with 1–>10 years of experience managing asthma patients (1 with <5 years, 2 with 5–10 years, and 3 with >10 years). All participants were aged between 30 and 50 years.

Generally, ED doctors expressed positive views of the clinical decision aids content and design. They found the information and flow clear, easy to follow and helpful. Specifically, they found the flow chart and hyperlinks easy to use.

Key refinements to the interactive decision aid included:

1. Addition of a warning box about not prescribing salbutamol when initiating a MART inhaler
2. Addition of spacer images alongside recommended inhalers
3. Integration of QR code with weblinks to inhaler technique videos
4. Improved heading and navigation to simplify use in time pressured settings

*HCP Training development*: Initial feedback from HCPs suggested that the training should be shorter and include pre-recorded materials for use in handovers. To improve accessibility, a 5-minute video alongside 30-minute in-person teaching sessions were developed. Feedback was obtained from 15 participants following the in-person teaching sessions delivered to ED doctors and ACPs after the decision aid had been introduced into clinical practice. HCPs found the training valuable for understanding the rationale for the change to MART prescribing, benefits of dry powdered inhalers (DPIs) and how to assess patient suitability, use of the decision aid and patient resources to support decision making and clarity of the prescribing guidelines and discharge documentation. Participants reported that the training and supporting materials were easy to follow and useful in time-limited ED settings.

HCP feedback identified key implementation challenges, including time pressures, competing ED demands, and limited availability of MART inhalers in discharge “to take away” (TTA) cupboards. Access to inhalers at discharge was viewed as important for in-person demonstration. Concerns were raised about additional workload related to adherence support, inhaler technique, documentation, and patient engagement, in a stretched service. The need for accessible patient information was highlighted. These insights directly informed the implementation package, underscoring the importance of streamlined processes, consistent stock availability, and accessible patient resources.

Key feedback, proposed modifications, rationale, and final agreed changes (with MoSCoW prioritisation) are summarised in Table 3. This transparent process demonstrates how patient and clinician feedback directly informed the final AMEND implementation package.

Using the TIDieR framework (32) we created a broad outline of the implementation package that included the content delivered, to whom and by whom, why, by what mode of delivery and how often, see Table 4.

**Table 4:**
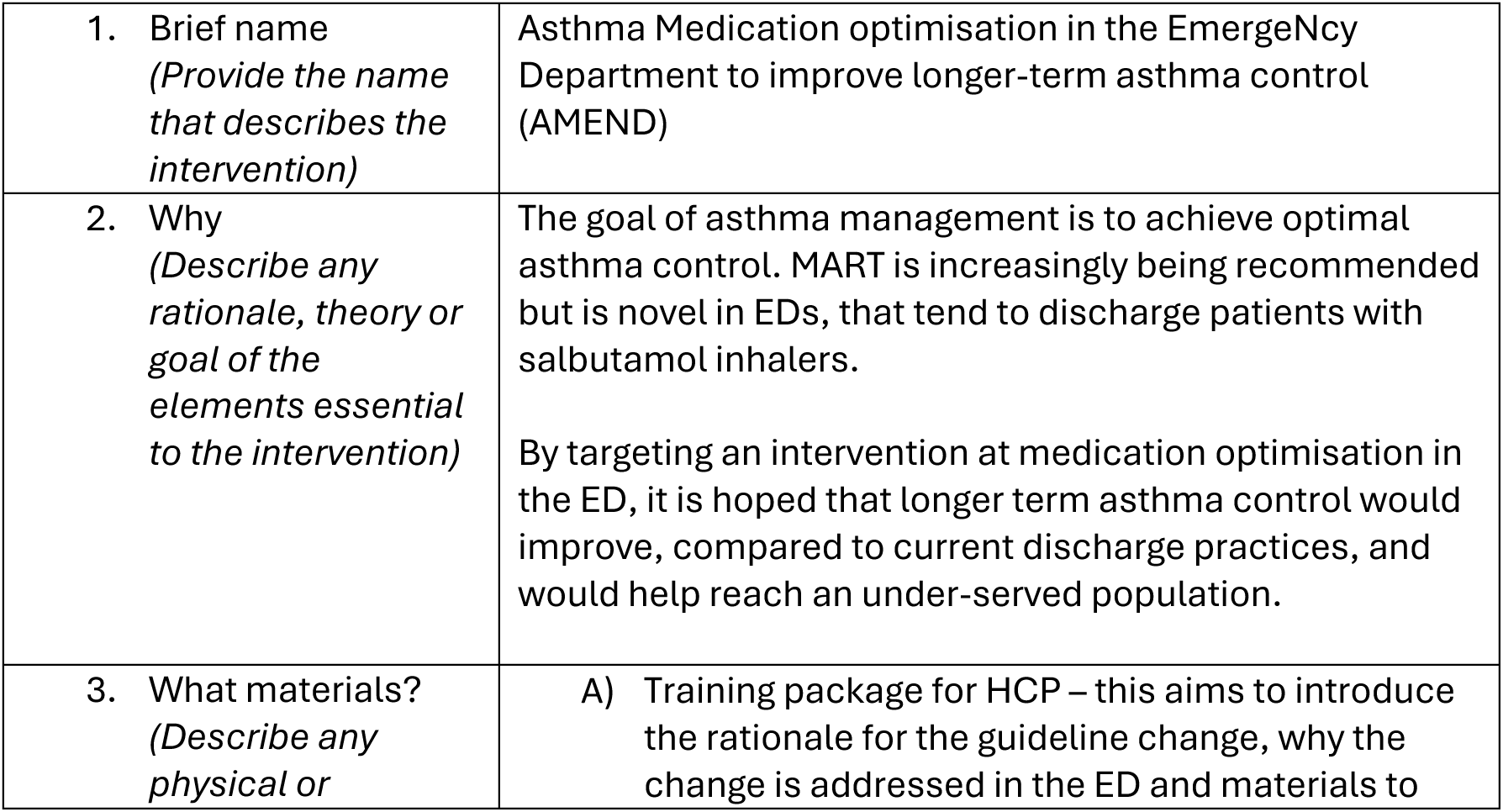
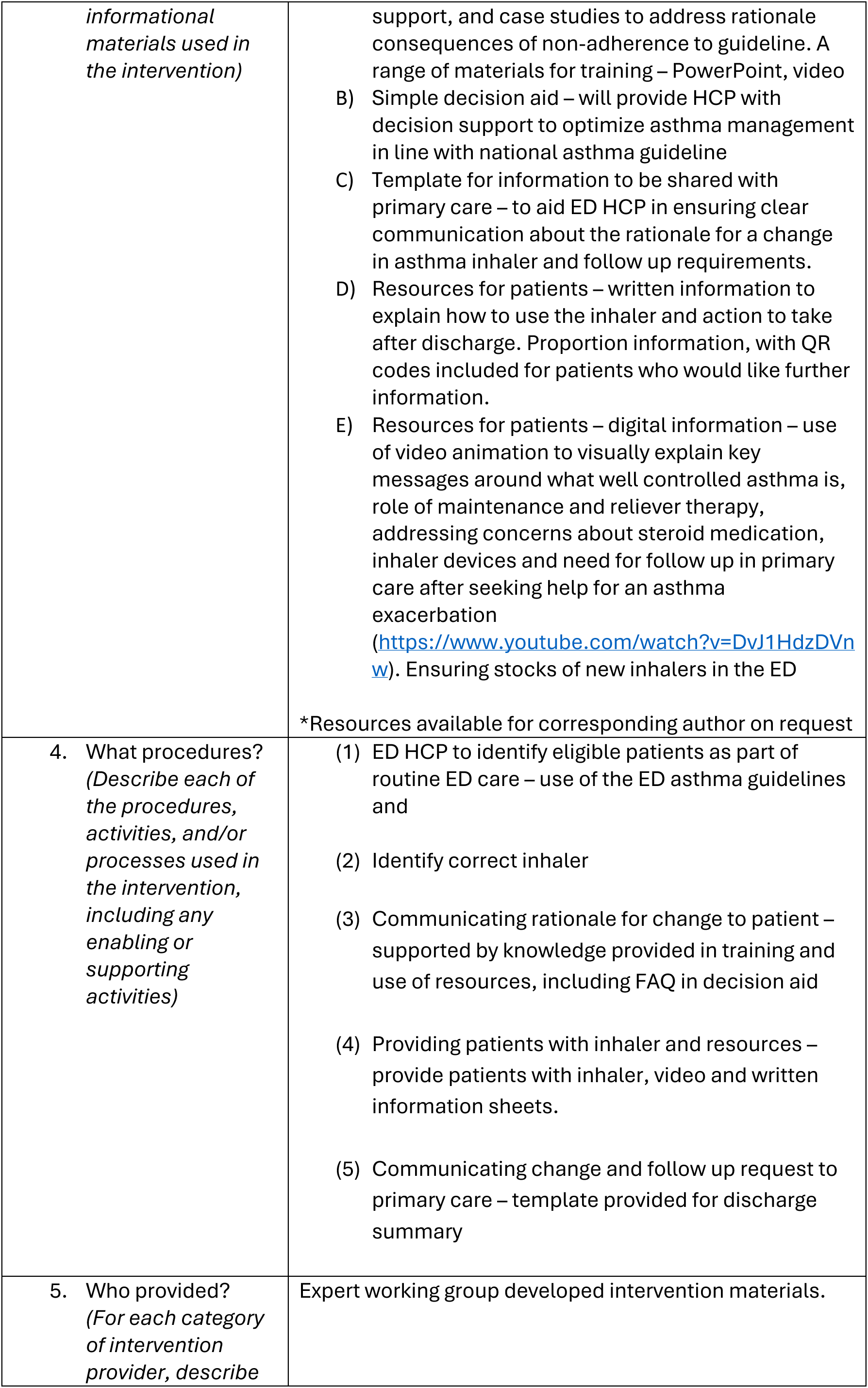
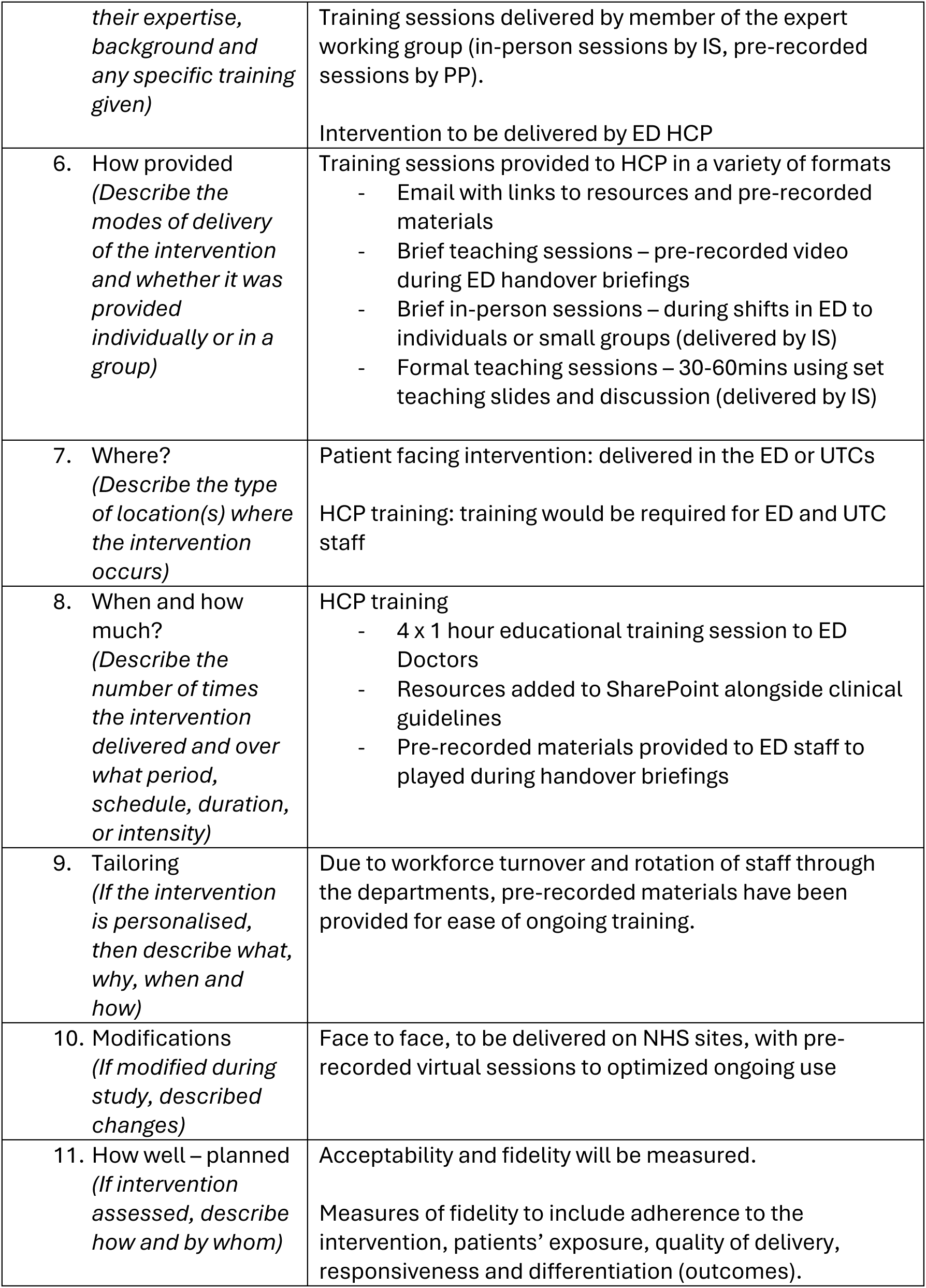
TIDier description of AMEND intervention.

|  |  |
| --- | --- |
| 1. Brief name<br>(Provide the name that describes the intervention) | Asthma Medication optimisation in the Emergency Department to improve longer-term asthma control (AMEND) |
| 2. Why<br>(Describe any rationale, theory or goal of the elements essential to the intervention) | The goal of asthma management is to achieve optimal asthma control. MART is increasingly being recommended but is novel in EDs, that tend to discharge patients with salbutamol inhalers.<br>By targeting an intervention at medication optimisation in the ED, it is hoped that longer term asthma control would improve, compared to current discharge practices, and would help reach an under-served population. |
| 3. What materials?<br>(Describe any physical or) | A) Training package for HCP – this aims to introduce the rationale for the guideline change, why the change is addressed in the ED and materials to |
| *informational materials used in the intervention)* | support, and case studies to address rationale consequences of non-adherence to guideline. A range of materials for training – PowerPoint, video<br>B) Simple decision aid – will provide HCP with decision support to optimize asthma management in line with national asthma guideline<br>C) Template for information to be shared with primary care – to aid ED HCP in ensuring clear communication about the rationale for a change in asthma inhaler and follow up requirements.<br>D) Resources for patients – written information to explain how to use the inhaler and action to take after discharge. Proportion information, with QR codes included for patients who would like further information.<br>E) Resources for patients – digital information – use of video animation to visually explain key messages around what well controlled asthma is, role of maintenance and reliever therapy, addressing concerns about steroid medication, inhaler devices and need for follow up in primary care after seeking help for an asthma exacerbation<br>(https://www.youtube.com/watch?v=DvJ1HdzDVnw). Ensuring stocks of new inhalers in the ED<br>*Resources available for corresponding author on request |
| 4. What procedures?<br>(Describe each of the procedures, activities, and/or processes used in the intervention, including any enabling or supporting activities) | (1) ED HCP to identify eligible patients as part of routine ED care – use of the ED asthma guidelines and<br>(2) Identify correct inhaler<br>(3) Communicating rationale for change to patient – supported by knowledge provided in training and use of resources, including FAQ in decision aid<br>(4) Providing patients with inhaler and resources – provide patients with inhaler, video and written information sheets.<br>(5) Communicating change and follow up request to primary care – template provided for discharge summary |
| 5. Who provided?<br>(For each category of intervention provider, describe | Expert working group developed intervention materials. |
| *their expertise, background and any specific training given)* | Training sessions delivered by member of the expert working group (in-person sessions by IS, pre-recorded sessions by PP).<br>Intervention to be delivered by ED HCP |
| 6. How provided<br>(Describe the modes of delivery of the intervention and whether it was provided individually or in a group) | Training sessions provided to HCP in a variety of formats<br>- Email with links to resources and pre-recorded materials<br>- Brief teaching sessions – pre-recorded video during ED handover briefings<br>- Brief in-person sessions – during shifts in ED to individuals or small groups (delivered by IS)<br>- Formal teaching sessions – 30-60mins using set teaching slides and discussion (delivered by IS) |
| 7. Where?<br>(Describe the type of location(s) where the intervention occurs) | Patient facing intervention: delivered in the ED or UTCs<br>HCP training: training would be required for ED and UTC staff |
| 8. When and how much?<br>(Describe the number of times the intervention delivered and over what period, schedule, duration, or intensity) | HCP training<br>- 4 x 1 hour educational training session to ED Doctors<br>- Resources added to SharePoint alongside clinical guidelines<br>- Pre-recorded materials provided to ED staff to played during handover briefings |
| 9. Tailoring<br>(If the intervention is personalised, then describe what, why, when and how) | Due to workforce turnover and rotation of staff through the departments, pre-recorded materials have been provided for ease of ongoing training. |
| 10. Modifications<br>(If modified during study, described changes) | Face to face, to be delivered on NHS sites, with pre-recorded virtual sessions to optimized ongoing use |
| 11. How well – planned<br>(If intervention assessed, describe how and by whom) | Acceptability and fidelity will be measured.<br>Measures of fidelity to include adherence to the intervention, patients' exposure, quality of delivery, responsiveness and differentiation (outcomes). |

These findings informed the final AMEND package which comprises five main components designed to be implemented as a whole: (1) a short animation video for patients (33) (2) written resources for patients (3) prescribing decision aid for HCP (4) teaching materials for HCP (5) template discharge text as described in Figure 4.

**Figure 4:**
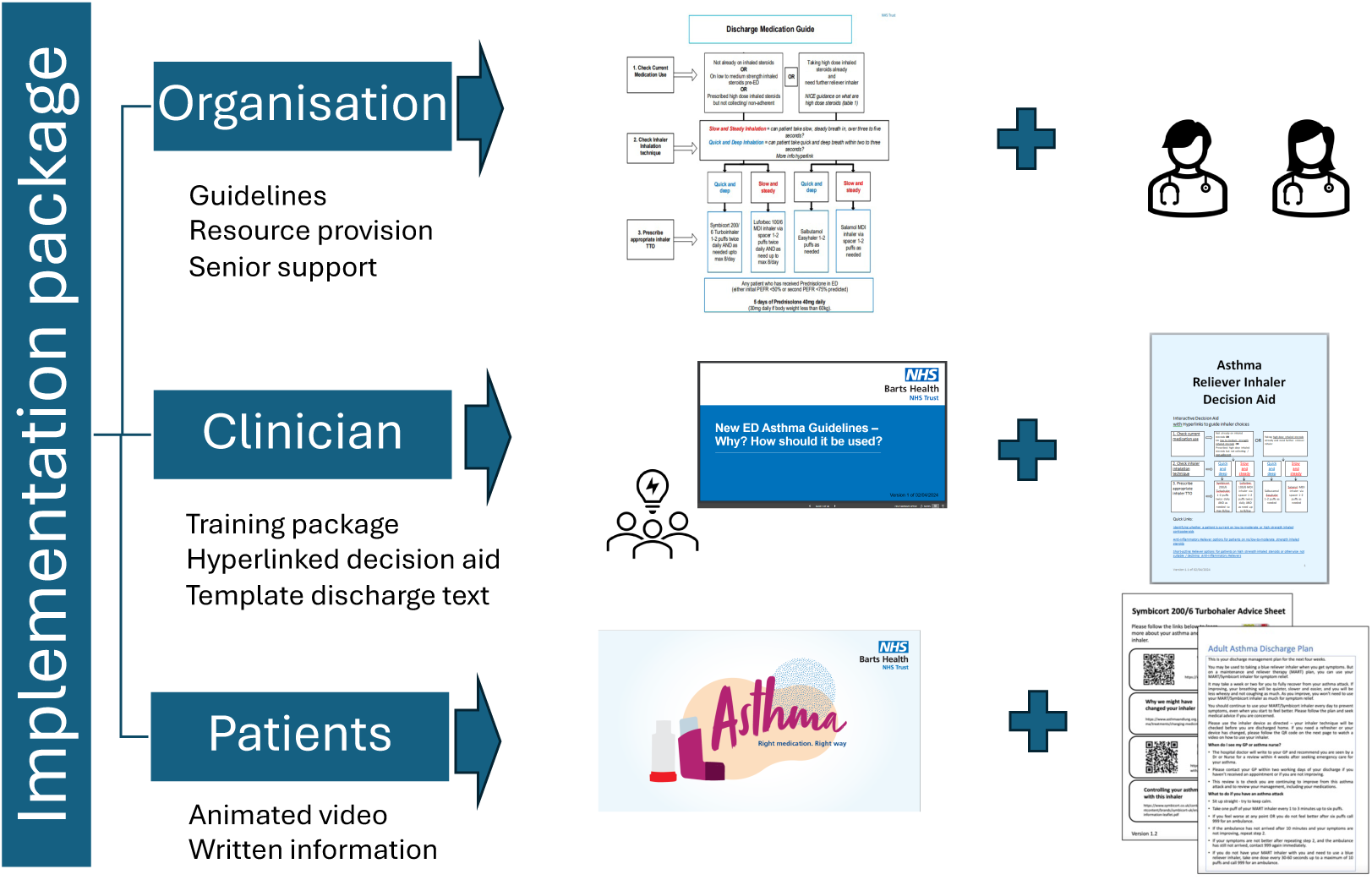
Summary of AMEND implementation package

## DISCUSSION

This paper describes the systematic development of AMEND, a theory informed implementation package designed to optimise asthma medication management - specifically initiation of MART- for adults presenting to the ED with uncontrolled asthma. AMEND supports the delivery of the updated NICE/BTS/SIGN Asthma guidelines (3) and targets ED discharge as a critical opportunity to promote supported self-management in a setting where health inequalities are often magnified. Developed using a multi-method approach, combining behavioural theory, qualitative research, and stakeholder engagement, AMEND adopts a whole systems perspective to identify modifiable behaviours, and practical constraints relevant to ED care (18, 34, 35). The resulting package supports non-respiratory specialists to initiate guideline concordant therapy, at ED discharge via a brief training package and clinical decision aid, aligned with national and international guidance (3, 36). Transparent reporting of this process provides a pragmatic model for intervention development in acute care settings.

Existing literature on asthma care in the ED has predominantly focused on patient education, with limited attention to changing long-term preventer therapy at the point of ED discharge (8). Patients generally prefer doctors to recommend a change in treatment and provide a rationale, and qualitative studies have suggested patients trust and would defer to such recommendation (15) (37).

Patients and healthcare professionals recognised the value of optimising preventer therapy at ED discharge, but reported barriers including time pressures, role boundaries, and uncertainty regarding prescribing guidelines (15, 16). Although decision support interventions can improve patient outcomes (38), clinician facing tools in electronic health records often suffer from poor uptake due to integration and engagement issues (39). These findings suggest that effectiveness alone is insufficient, tools must be designed for usability and acceptability in high-pressure ED settings. AMEND was designed as a simple, low burden flow diagram with embedded links for further information, aligning with existing practices, rather than requiring changes to electronic systems. Supporting HCP confidence in recommending medication changes was central to the intervention (16), with resources facilitating consistent, evidence-based messaging, and proportionate written and digital information to support self-management after discharge.

Visual and patient facing resources were included to reinforce teachable moment during ED visits, facilitating behaviour change, such as medication optimisation (40, 41). ED attendance has been associated with patients following treatment advice (7) highlighting this as a key opportunity to influence long-term outcomes. Brief visual interventions, such as bedside animations, have improved treatment control perceptions and reducing medication harm (42), while pictures of inhalers and mechanisms aid patients with language barriers or literacy barriers (43). AMEND incorporates both an animation video and written information sheets with embedded links to reinforce key messages and support self-management addressing the common problem of poor recall of information following ED visits.

Organisational factors and sustainability were also considered. The package was designed to integrate with existing ED workflows, requiring minimal additional resources, with senior HCP support highlighted as key (44).These design choices increased the likelihood of sustained use and potential replication in other ED settings.

### Innovations and Contributions

The development of AMEND also reinforces previous ED research (23, 45, 46, 47, 48, 49, 50, 51) in finding that the TDF is useful for identifying important domains for change behaviour interventions. In the ED context, knowledge, environmental context and resources and professional role (47, 48, 50, 51). AMEND demonstrates that behavioural frameworks can guide intervention design even in complex high intensity settings. AMEND is among the first implementation packages to apply behavioural frameworks systematically to support prescribing behaviours in the ED. Using a PBA approach, allowed us to accommodate diverse stakeholder perspectives (11) resulting in a multifaceted, acceptable package. The development process was transparent and systematic, linking behavioural theory to intervention content through a logic model and guiding principles, highlighting both the value of optimisation for this population and the potential for replication in other acute care contexts.

### Practical Implications - Impact on Healthcare Practice

The AMEND implementation package has been designed for easy integration into routine ED practices, to support safe and effective initiation of guideline-concordant preventer therapy, such as MART, during acute presentations, thereby supporting faster implementation of guidelines in clinical practice. The decision aid can be embedded into local prescribing guidelines, accessible via the intranet and used during clinical decision making without disrupting time sensitive care. It also functions as a training tool, supporting prescriber confidence in initiating MART regimes in non-specialist settings and utilising a teachable and reachable moment for groups such as young adults who often tolerate symptoms without seeking preventative help (52). While new NICE/BTS/SIGN guidelines are an important starting point to increase knowledge and awareness, alone they are unlikely to change behaviour of the clinical teams in EDs. Evidence internationally suggests that the new MART regime is being poorly implemented in the outpatient setting, with many patients still being given a SABA reliever inhaler (53). Multiple complex behaviours are required across the system to implement such guidelines and support adults optimally. AMEND has potential to be an important facilitator of these.

For patients, the tailored materials offer clear, actionable explanations and discharge instructions that support medication adherence beyond the ED visit. As such, AMEND has the potential to reduce repeat attendances, and promote safer asthma care in emergency settings.

There is potential to expand this implementation package to a wider group of asthma patients presenting to unscheduled care with their asthma including GPs, urgent treatment centres and on discharge from hospitals wards; the benefits in these contexts should be tested. The implementation package has been designed with considerations for a diverse demographic base, but further tailoring with considerations for cultural competence, language and health literacy may still be required.

### Strengths and Limitations

Key strengths include the rigorous development informed by theory, evidenced and multidisciplinary input, including patient and public involvement. Limitations included a phased roll out during updates to national guidance which may impact on the HCP beliefs and behaviours and impact the emphases of key messages within the training materials.

Initiating a new medication regime and discontinuing an existing one are not equivalent behaviours. While the AMEND implementation package was designed to support initiation of MART and optimise adherence, it did not explicitly target the discontinuation of salbutamol overuse. Nevertheless, given the interconnected nature of prescribing and adherence behaviours, we hope the resources developed might also contribute indirectly to reductions in inappropriate salbutamol use. Future work should consider strategies that more explicitly address discontinuation behaviours alongside initiation.

Future work should assess implementation, clinical outcomes and equitable impact across demographic groups, with potential refinement of delivery modes and engagement of broader professional groups (i.e. nursing and pharmacy professionals who play a key role in discharge and education).

## Conclusion

AMEND provides a theory informed, pragmatic implementation package to support the optimisation of asthma medication at ED discharge. It addresses known barriers to change, aligns with guideline recommendations, and offers transferable learning to implementing complex interventions in high pressure, acute care settings. A future evaluation is proposed to assess both implementation and clinical outcomes.

## List of abbreviations

ACP: Advanced Care Practitioner
AMEND: Asthma Medication Optimisation in the Emergency Department
APEASE: Acceptability, Practicability, Effectiveness, Affordability, Side-effects and Equity
AUKCAR: Asthma UK Centre for Applied Research
BCW: Behaviour change wheel
BCT: Behaviour change technique
BTS: British Thoracic Society
CSM: Common-sense model
DPI Dry: Powered Inhaler
ED: Emergency Department
FY: Foundation Year
GINA: Global Initiative for Asthma
GP: General Practitioner
HCP: Health care professional
HRA: Health Research Authority
ICS: Inhaled corticosteroids
MART: Maintenance and Reliever Therapy
MRC: Medical Research Council
NCF: Necessity Concerns Framework
NHS: National Health Service
NICE: National Institute for Health and Care Excellence
PAPA: Perceptions and practicalities approach
PBA: Person Based Approach
PPI: Patient and Public Involvement
REC: Research Ethics Committee
SABA: Short acting beta-agonist
ST: Speciality Trainee
TDF: Theoretical Domains Framework
TTA: To Take Away
UK: United Kingdom

---

**Additional file 1: TIDier description of AMEND intervention**

## DECLARATIONS

### Human Ethics and Consent to Participate

Ethical approval for the qualitative research in phase 1 and 2 was obtained from Camden and Kings Cross Research Ethics Committee HRA Approval [REC Reference 21/LO/0665]. The study received NHS governance approval from Barts Health NHS Trust. All participants provided written informed consent.

Phase 3, including the user testing and refinement were conducted as part of a quality improvement project which was registered at Barts Health to design the implementation package, implement and evaluate the acceptability. This did not require separate ethical approval.

Stakeholders were informed that participation was voluntary, that they could withdraw at any time and that all data would be treated confidentially. Verbal informed consent was obtained from all participants in the think aloud interviews prior to participation. For online workshops and structured feedback via Microsoft Forms, participation was voluntary and consent was by participation.

### Declaration of competing interests

IS has given lectures at meetings with honoraria supported by AstraZeneca, is conducting quality improvement activity at their institution supported by AstraZeneca. IS received a PhD studentship from AUKCAR which supported the initial work including the systematic review and data collection for the interviews from patients and healthcare professionals.

BB is conducting quality improvement activity at their institution supported by AstraZeneca. KP has given lectures at meetings/webinars, with/without honoraria, supported by Sanofi; has taken part in clinical trials sponsored by AstraZeneca.

PEP has attended advisory boards for AstraZeneca, GlaxoSmithKline and Sanofi; has given lectures at meetings/webinars, with/without honoraria, supported by AstraZeneca, Chiesi and GlaxoSmithKline; has attended international conferences with AstraZeneca; has taken part in clinical trials sponsored by AstraZeneca, GlaxoSmithKline, Novartis, Regeneron and Sanofi; is conducting research funded by GlaxoSmithKline for which his institution receives remuneration and quality improvement activity at his institution supported by AstraZeneca.

### Funding

The AMEND quality improvement project was funded by AstraZeneca through an Education Grant and AstraZeneca was not involved in the development of the programme educational materials. The views expressed are those of the authors and not necessarily those of AstraZeneca. The AMEND project builds upon IS’s PhD studentship from AUKCAR, which was funded by Asthma+Lung UK as part of the Asthma UK Centre for Applied Research [AUK-AC-2012-01 and AUK-AC-2018-01].

### Contributors

The development of this implementation package was conceptualised by IS with support from PP and LS. It followed on from IS’s PhD project, who was supervised by LS, PP, KP & CG. IS, BB, JB, ADS, PP and LS drafted and refined the implementation package materials with feedback from all authors. IS conducted the data analyses with support from PP and LS and with feedback from all authors. IS drafted the initial manuscript, with PP and LS substantively contributing to revisions. All authors critically reviewed the manuscript, contributing important intellectual content and approved the final manuscript. PP and LS share joint senior authorship.

### Consent for publication

Not applicable

### Availability of data and materials

The datasets used and/or analysed during the current study are available from the corresponding author on reasonable request.

The materials in the AMEND implementation package are available from the corresponding author on reasonable request

### Patient and public involvement

Patients and/or the public were involved in the design, or conduct, or reporting, or dissemination plans of this research. Refer to the Intervention development methodology section for further details.

## Data Availability

The datasets used and/or analysed during the current study are available from the corresponding author on reasonable request.
The materials in the AMEND implementation package are available from the corresponding author on reasonable request

